# The Genetic Architecture of Self-Reported Psychopathy in Adults

**DOI:** 10.64898/2026.09.08.26362549

**Authors:** Justin D. Tubbs, Travis T. Mallard, Maria Dalby, Younga H. Lee, Karmel W. Choi, Tian Ge, Niels Plath, Lene Hammer-Helmich, Julie M. Granka, 23andMe Research Team, Essi Viding, David A. Hinds, Jordan W. Smoller, Joshua W. Buckholtz

**Affiliations:** Psychiatric and Neurodevelopmental Genetics Unit, Center for Genomic Medicine, Massachusetts General Hospital, Boston, MA; Center for Precision Psychiatry, Department of Psychiatry, Massachusetts General Hospital, Boston, MA; Department of Psychiatry, Harvard Medical School, Boston, MA; Stanley Center for Psychiatric Research, Broad Institute of MIT and Harvard, Cambridge, MA; H. Lundbeck A/S, Valby, Denmark; 23andMe Research Institute, Palo Alto, CA, USA; Division of Psychology and Language Sciences, University College London, London, U.K; Department of Psychology, Stanford University, Stanford, CA

## Abstract

**Background:** Psychopathy is a personality construct characterized by affective and interpersonal features coupled with severe antisocial behavior. While strongly associated with extreme behaviors such as violent crime, empirical work suggests that psychopathy exists as a dimensional trait in the general population. However, our understanding of the genetic structure of psychopathy remains limited, with genome-wide association studies (GWAS) having been restricted to broad measures of antisocial behavior.

**Methods:** We perform the first large-scale GWAS of psychopathy, using total and factor scores from the Self-Report Psychopathy scale (SRP) in up to 10,558 participants enriched for mood disorder cases from the AFFECT study. LD score regression (LDSC) was used to characterize the heritability of SRP scores and estimate their genetic correlation (rG) with several externally-measured traits, using the Benjamini-Hochberg approach to perform multiple testing correction.

**Results:** Psychopathy scores showed significant heritability, with 12-14% of phenotypic variance explained by common genetic variants. However, GWAS did not identify any genome-wide significant loci in this sample. The LDSC-estimated genetic correlation with antisocial behavior was only 0.54 (95%CI: 0.21-0.81), in line with prior twin research. Significant genetic correlations were found with a wide range of psychiatric disorders (rG’s: 0.21 - 0.72), with the highest observed for borderline personality disorder (95% CI: 0.41-0.72) and substance use disorders (95% CI: 0.43-0.63). Psychopathy was also genetically correlated with lower life-satisfaction, poorer health outcomes, worse cognitive functioning, and lower educational attainment.

**Conclusions:** In the first large-scale GWAS of self-reported psychopathy, we found significant SNP heritability and genetic overlap with risk for psychiatric disorders, and poorer behavioral, cognitive, and physical health outcomes. These findings provide important genetic support for differentiating psychopathy from antisocial behavior. Further, they inform our understanding of how genetic factors may contribute to poor health outcomes in psychopathy, and shed insight into the comorbidity between psychopathy and other disorders.

## Introduction

Psychopathy is characterized by the combination of emotional-interpersonal deficits and extreme antisocial behavior^1^. Individuals with psychopathy are often described as superficially charming, possessed of a shallow emotional experience, and lacking in remorse, guilt and regret^1–4^. Psychopathy is strongly associated with criminal behavior, recidivism, and poor treatment outcomes^5–8^, and psychopathy scores are increasingly used in legal contexts to inform decisions about parole and civil commitment^9–13^. Individuals with psychopathy impose profound costs to society due to the direct and indirect expenses associated with incarceration, treatment, opportunity costs and lost productivity associated with crime^14,15^. Moreover, psychopathy is associated with significant morbidity and mortality; in addition to lower life expectancy, higher scores on measures of psychopathy are linked to reductions in overall health and increased risk for a range of chronic health conditions^16–18^. Despite its impact to affected individuals and to society as a whole, relatively little is known about the etiology of psychopathy.

Psychopathy can be conceptualized as a unique cluster of personality and behavioral traits situated within the broader domain of antisocial and externalizing psychopathology^1,19–22^. In addition to the presence of antisocial lifestyle traits, psychopathy is distinguished from antisocial personality disorder (ASPD) and externalizing by prominent trait-like affective–interpersonal features that are neither necessary nor sufficient for ASPD diagnoses or for high scores on broadband externalizing measures. Classic and contemporary formulations of psychopathy emphasize callous–unemotional traits, shallow or constricted affect, lack of empathy and remorse, manipulativeness, and bold, socially dominant behavior alongside chronic rule-breaking. By contrast, ASPD and related externalizing constructs are defined primarily by behavioral deviance—impulsivity, irresponsibility, substance abuse, aggression, and persistent violation of social norms—with considerably less weight placed on the emotional–interpersonal features that typify psychopathy^1,2,4,23–25^. As a result, most individuals with high levels of psychopathy would also meet criteria for ASPD or score high on externalizing measures; yet, the converse is not true: despite exhibiting persistent antisocial behavior, most individuals who meet criteria for ASPD or with high externalizing factor scores would not necessarily show high levels of psychopathy^1,26^. Dimensional work on the externalizing spectrum further suggests that the relationship between psychopathy and ASPD/externalizing is driven by the social deviance and antisocial lifestyle features of the former. Affective–interpersonal features appear to capture variance that is only partially shared with broadband externalizing and may have distinct neural and genetic underpinnings^2,4,27–30^.

Twin and family studies indicate that broadband antisocial behavior and externalizing psychopathology are moderately heritable, with meta-analytic estimates suggesting that roughly 40% of the variance in antisocial behavior reflects genetic factors (additive heritability [*h*²] ≈ 0.32, plus additional non-additive genetic effects), alongside smaller but non-trivial shared environmental influences^29^. Quantitative genetic work using latent externalizing factors—capturing the shared liability across conduct problems, antisocial personality disorder, and substance use disorders—further suggests that this common externalizing dimension is strongly heritable (often >60%), and is consistent with a genetic liability for transdiagnostic disinhibitory psychopathology^27,31–34^. By contrast, DNA-based estimates from GWAS are considerably smaller: the initial Broad Antisocial Behavior Consortium analysis (N ≈ 16,000) reported SNP heritability (*h^2^_snp_*) of ∼5% for broadly defined antisocial behavior and identified no= robust genome-wide significant loci, underscoring a highly polygenic architecture and substantial “missing heritability” relative to twin-based estimates^35^. A subsequent meta-analytic GWAS of broad antisocial behavior (N ≈ 85,000) yielded several genome-wide significant loci and a SNP heritability estimate around 7%, along with polygenic scores that modestly predict antisocial outcomes across independent cohorts^36^. Multivariate genomic approaches that treat externalizing as a latent factor across antisocial behavior, ADHD, and substance use have substantially increased power: the Externalizing Consortium’s EXT GWAS in ∼1.5 million individuals identified hundreds of loci associated with a broad externalizing factor and produced polygenic scores that explain up to ∼10% of the variance in externalizing-related outcomes, highlighting a diffuse, pleiotropic architecture spanning self-regulation and reward-related traits^37,38^.

Converging evidence from twin and genomic studies indicates that individual differences in psychopathic traits are substantially heritable^1,39–41^. Further, work in genetically informative samples suggests potentially differential heritability for antisocial behaviour depending on whether it is accompanied by affective psychopathic traits or not (e.g. heritability estimates of 81% vs. 30%)^39^, though this research has been limited to youth and adolescents^29^. Together, these results implicate strong genetic liability to psychopathic personality from childhood. Recent reviews of the literature confirm that psychopathic traits, including both interpersonal–affective and behavioral facets, consistently show modest-to-high heritability, with the largest estimates typically observed for CU traits^29,42^. This points to the possibility that psychopathic traits have a genetic architecture that is at least partially unique with respect to antisocial behavior *per se.* To date, little is known about the common-variant architecture of psychopathy, and the degree to which liability for psychopathy is shared with that for antisocial behavior, psychiatric disorders, and measures of health and health-related behaviors, personality, well-being, cognition and education.

Psychopathy and broadband antisocial behavior show significantly overlapping, but not identical, patterns of health burden. This may be due, in part, to distinct patterns of cognitive^43,44^ and neurobiological^45,46^ impairment. Externalizing – a dimensional latent construct indexing liability for substance abuse, antisocial behavior and impulsive traits – is linked to a diffuse pattern of comorbidity, including increased risk for personality disorders, as well as mood and anxiety disorder symptoms and diagnoses^27,47–49^. Psychopathy is also associated with increased risk for personality disorders, impulsive traits and conditions such as attention deficit/hyperactivity disorder, and individuals high in psychopathy exhibit elevated rates of substance use disorder^1,50,51^. These phenotypic associations appear relatively selective for the antisocial/lifestyle features of psychopathy^1,51^. Interestingly, associations between psychopathy and internalizing psychopathology appear to be factor specific: while moderate positive correlations between internalizing and the antisocial/lifestyle features of psychopathy have been reported, some have observed lower trait anxiety in individuals with higher scores on measures of psychopathy’s interpersonal/affective features^1,51,52^. Intriguingly, one study in community adolescent twins used the Multidimensional Personality Questionnaire (MPQ) to assess psychopathic traits and their genetic relationships to externalizing and internalizing psychopathology. MPQ-assessed “fearless dominance” (analogous to the interpersonal/affective features indexed by other measures of psychopathy) was associated with reduced risk for internalizing^53^. While recent molecular genetic work has begun to identify the degree to which phenotypic associations with broadband antisocial behavior and related constructs, such as self-control^54^ are driven by genetic factors^35–37,55^, the genetic underpinnings of psychopathy’s network of comorbidity remains largely unexplored. The purpose of the present study is to bridge the gaps in our current understanding of the genetic architecture of psychopathy by performing the first GWAS of self-reported psychopathic traits in an adult sample.

## Methods

### Sample

Data were collected as part of the AFFECT study^56^, a longitudinal cohort designed to examine genetic associations with cognition and symptom domains in a sample enriched for mood disorders. A detailed description of the study design is provided in a prior publication^56^. Briefly, 48,467 participants were recruited through the 23andMe Research Institute participant database and through social media, including 14,768 cases with a history of major depression, 9,864 with a history of bipolar disorder, and 23,835 controls without a history of either. Mood disorder status was indicated through self-reported diagnosis by a medical professional. Participants were asked to complete a set of monthly online assessments over the course of nine months which included demographics, medication information, symptom scales, and cognitive batteries. Participants provided informed consent and volunteered to participate in the research online, under a protocol approved by the external AAHRPP-accredited IRB, Ethical & Independent (E&I) Review Services. As of 2022, E&I Review Services is part of Salus IRB (https://www.versiticlinicaltrials.org/salusirb).

### Phenotype Assertainment

The self-reported psychopathy short-form (SRP-SF)^57^ was administered at the eighth assessment and was completed by 28% of all participants, including 4,463 subjects with a history of major depression, 2,474 with a history of bipolar disorder, and 3,621 without a history of mood disorders. The SRP-SF is a well-validated measure designed to assess psychopathy in community samples^57,58^, and indexes four major facets: interpersonal, affective, lifestyle, and antisocial. For each participant with available data, we calculated a total (sum) score and two higher-order factor scores. Factor 1 (F1) includes interpersonal and affective components such as deception, manipulation, and lack of empathy or guilt. Factor 2 (F2) includes socially deviant lifestyle features such as impulsivity, sensation-seeking, behavioral disinhibition, and frank antisocial behavior (i.e. violence proneness, rule breaking preference, substance abuse, criminal behavior). Mood disorder cases had statistically significant, modestly higher log-transformed F1 (Cohen’s d = 0.21, p < 2.2e-16), F2 (Cohen’s d = 0.50, p < 2.2e-16), and Total SRP scores (Cohen’s d = 0.48, p < 2.2e-16).

### Genotyping

DNA was collected through saliva samples, with extraction and genotyping performed at the National Genetics Institute, a CLIA-licensed clinical laboratory under the Laboratory Corporation of America. SNP genotyping, imputation, and quality control were performed by 23andMe following a standardized protocol, as described in a prior publication^56^, resulting in approximately 9.2 million high-quality SNPs for GWAS analysis.

### Statistical Analyses

GWAS were performed by the 23andMe Research Institute for each of the three psychopathy scores. Specifically, linear regression was performed on the log-transformed scores for each SNP, with covariates for age, sex, the first five genetic principal components, and genotyping platform. GWAS included the maximally unrelated subset of participants^59^ with European-like ancestry. This resulted in 10,558 participants included in the GWAS of F1, 10,536 for F2, and 10,511 for the total psychopathy score. Summary statistics were subsequently filtered to exclude any variants with imputation INFO scores below 0.9, minor allele frequency less than 0.01, or not contained in the 1000 genomes reference panel. LD score regression (LDSC)^60,61^, implemented through the GenomicSEM R package^62^, was first used to estimate the SNP heritability (*h^2^_snp_*) of the three psychopathy scores. LDSC was also used to estimate the genetic correlation (rG) between psychopathy scores and 41 other traits using GWAS summary statistics from external studies (Supplementary Table 1), all of which exhibited statistically significant *h^2^_snp_*. These included 14 psychiatric disorders, 13 traits related to education and cognition, 8 traits related to personality and well-being, and 5 traits related to health and health behaviors. To account for multiple-testing across external traits, we employed a false-discovery rate (FDR) correction within the set of tests for each combination of domain and psychopathy score using the Benjamini-Hochberg method.

### Sensitivity Analyses

Given that cases and controls showed significant mean differences in SRP scores, we performed a sensitivity analysis to assess the potential impact of the inclusion of mood disorder cases in our analysis. Specifically, we used GenomicSEM to fit two nested linear regression models estimating the genetic association between the three SRP phenotypes and each of the 41 external traits. The first model estimated the marginal association between each SRP trait and each external trait, while the second model estimated the conditional association by including mood disorder case-control status as a covariate.

## Results

### Genome Wide Association

GWAS results for the total, F1, and F2 psychopathy scores are shown in Figure 1 (QQ plots are shown in Supplementary Figure 1). No genome-wide significant loci were identified for any of the three scores. However, LDSC indicated that all three phenotypes exhibited significant *h^2^_snp_* (Figure 2 diagonal entries, Supplementary Table 2). The *h^2^_snp_* for F1 was estimated to be 0.12 (SE=0.05, p = 0.009), while *h^2^_snp_* estimates for both F2 and the total score of 0.14 (SE=0.05, p=0.002). All three scores exhibited high genetic inter-correlations, which were not significantly different from 1. The genetic correlation between F1 and F2 was 0.86 (SE = 0.3), while the genetic correlation between the total score and F1 and F2 were 0.96 (SE = 0.34) and 0.97 (SE = 0.31), respectively. Notably, the SNP heritability estimate for psychopathy observed here (∼0.14) falls toward the upper end of the range reported for individual externalizing-related phenotypes, though direct comparisons to latent factor–level heritability estimates and are not straightforward given their dependence on model specification^36,37^.

**Figure 1.**
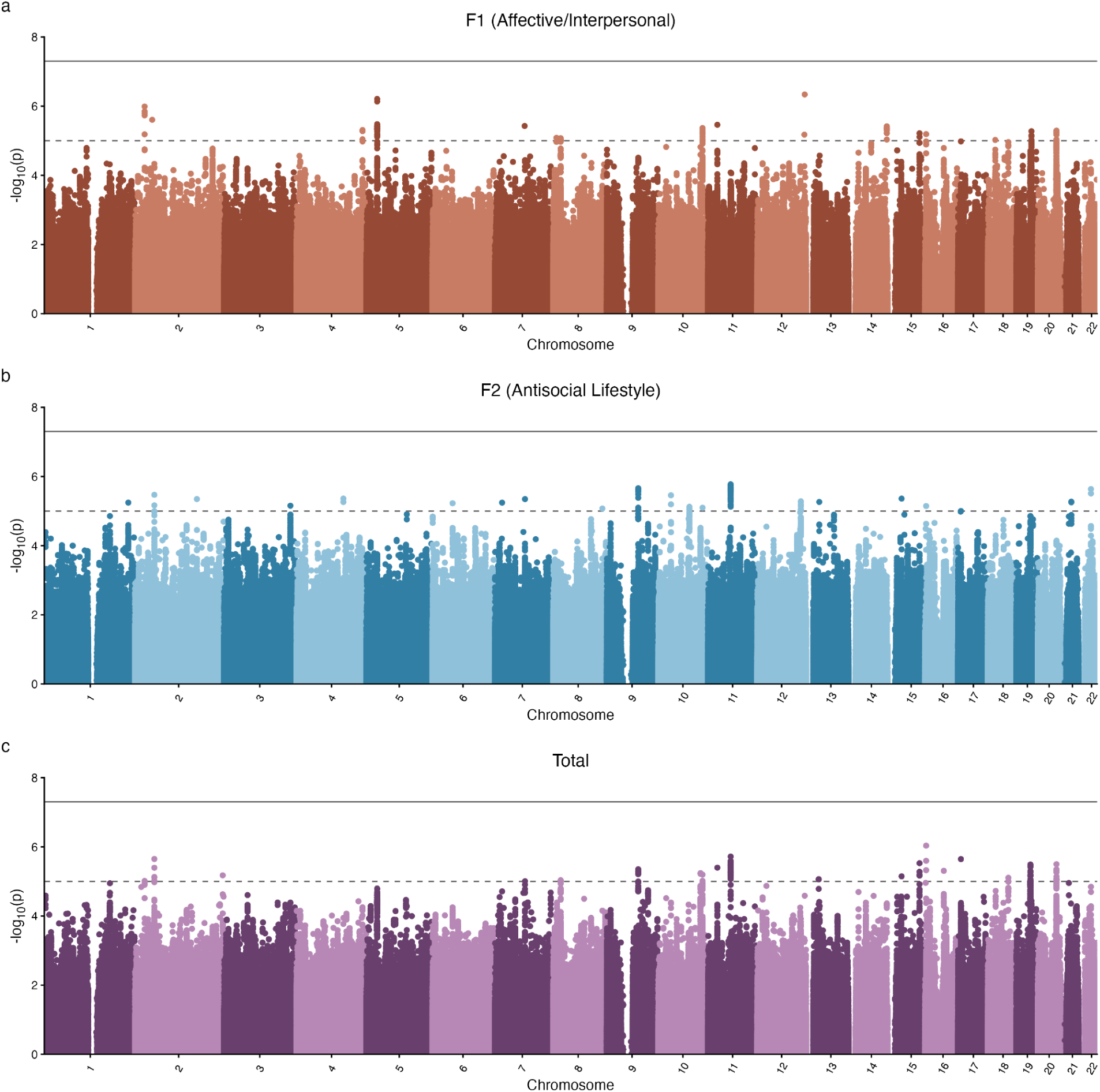
Polygenic architecture of self-reported psychopathy factors. Note. Manhattan plots for self-reported psychopathy (SRP) F1 (panel a), F2 (panel b), and total (panel c) scores. Points representing single nucleotide polymorphisms are ordered by chromosomal position on the x axis, with the y axis indicating the -log_10_p-value of association with the given log-transformed SRP score.

**Figure 2.**
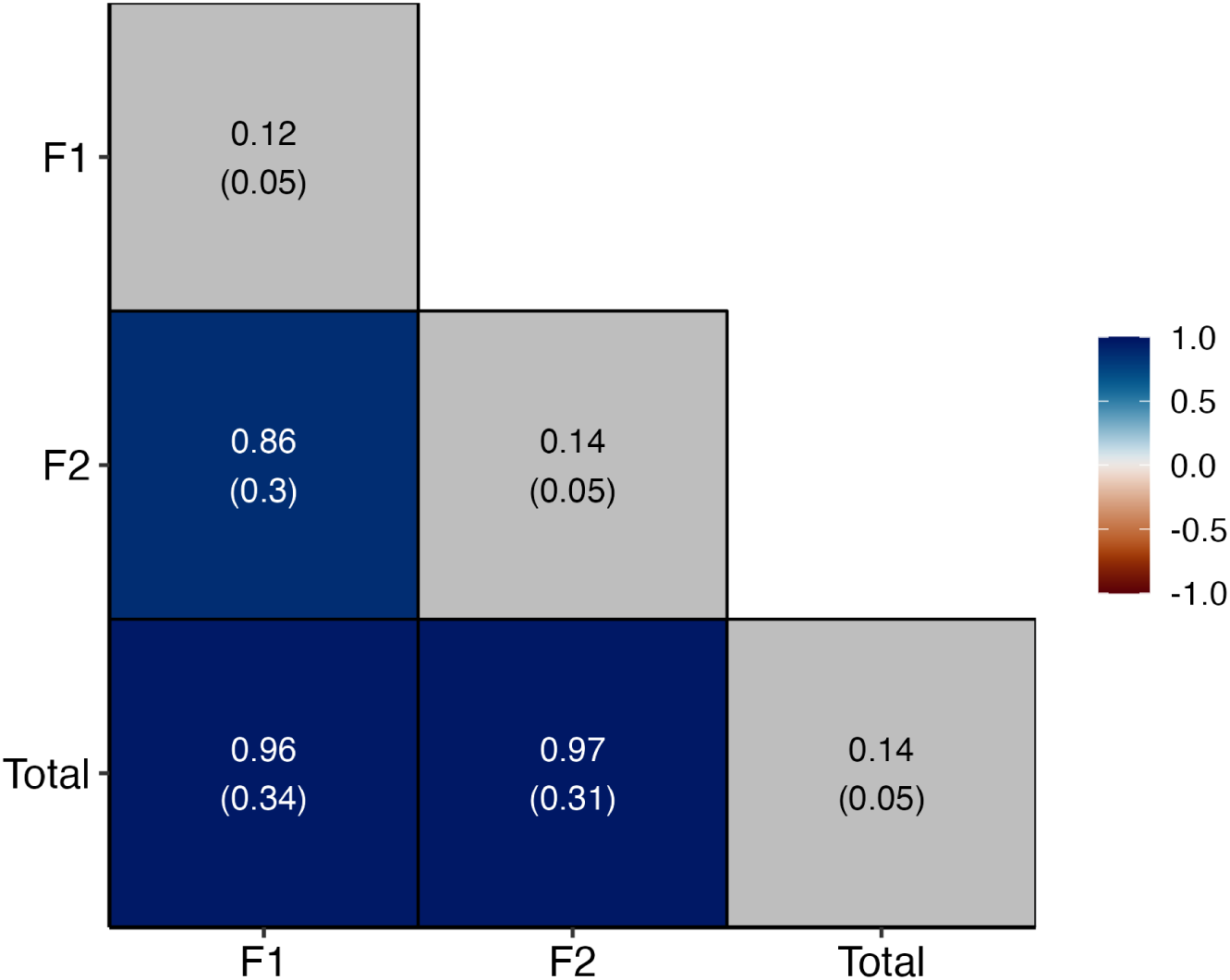
Self-reported psychopathy shows moderate SNP heritability and high genetic overlap between phenotypic factors. Note. A heatmap of genetic correlations between self-reported psychopathy F1, F2, and total scores. Off diagonal elements denote genetic correlations, while diagonal elements denote SNP heritabilities estimated using LD score regression. Standard errors are given in parentheses below each estimate.

### Genetic Correlations

#### Psychiatric Disorders

Psychopathy scores showed widespread and substantial genetic correlation with several psychiatric disorders (Figure 3, Supplementary Table 3). The total score was significantly genetically correlated with borderline personality disorder (rG = 0.59, SE = 0.10, P_FDR_ = 2.3e-8), attention-deficit hyperactivity disorder (rG = 0.48, SE = 0.08, P_FDR_ = 2.3e-8), major depressive disorder (rG = 0.43, SE = 0.11, P_FDR_ = 4.9e-4), post-traumatic stress disorder (rG = 0.41, SE = 0.17, P_FDR_ = 0.03), autism spectrum disorder (rG = 0.27, SE = 0.12, P_FDR_ = 0.03), and schizophrenia (rG = 0.20, SE = 0.07, P_FDR_ = 9.6e-3). For all three substance use disorders considered, we also found significant genetic correlations with the total score. Notably, genetic correlations with substance use disorders were nominally higher for F2 compared to F1 or total scores. This is consistent with the notion that F2 traits account for much of the overlap between psychopathy and externalizing. Psychopathy scores showed no significant genetic correlation with obsessive-compulsive disorder, Tourette’s syndrome, anxiety disorders, or anorexia nervosa, nor did we find evidence for a protective effect of F1 traits on mood disorders.

**Figure 3.**
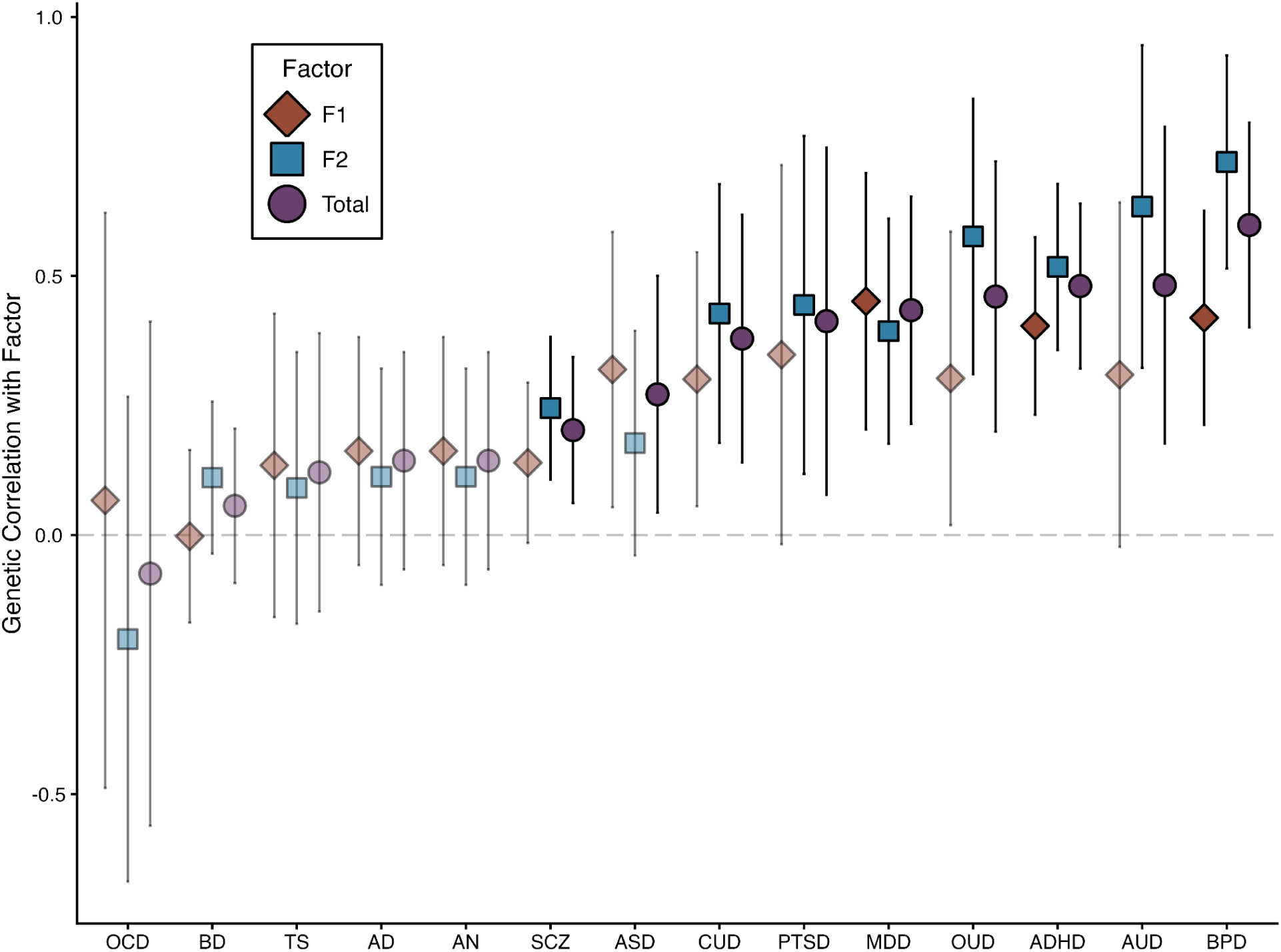
Self-reported psychopathy exhibits widespread genetic correlations with psychiatric disorder liability. Note. A forest plot of genetic correlations estimated using LD score regression between 14 psychiatric disorders and self-reported psychopathy F1 (in red diamonds), F2 (in blue squares), and total (in purple circles) scores. Estimates with false-discovery rate q-values less than 0.05 are opaque, while non-significant estimates are translucent. Error bars denote 95% confidence intervals around each estimate. ADHD = attention-deficit hyperactivity disorder, AN = anorexia nervosa, AD = anxiety disorders, ASD = autism spectrum disorder, AUD = alcohol use disorder, BD = bipolar disorder, BPD = borderline personality disorder, CUD = cannabis use disorder, MDD = major depressive disorder, OCD = obsessive compulsive disorder, OUD = opioid use disorder, PTSD = post-traumatic stress disorder, SCZ = schizophrenia, TS = Tourette’s syndrome.

#### Health-Related Behavioral Traits, Health Outcomes, and Social Determinants of Health

Psychopathy scores also exhibited several significant associations with health and health-related behavioral traits (Figure 4a). The total psychopathy score showed a significant positive genetic correlation with a previous GWAS of broad antisocial behavior (rG = 0.54, SE = 0.15, P_FDR_ = 6.0e-4), as well as a broad index of behavioral risk-taking (rG = 0.51, SE = 0.08, P_FDR_ = 2.9e-9). In addition, total psychopathy was also genetically correlated with poorer health outcomes including higher BMI (rG = 0.25, SE = 0.07, P_FDR_ = 4.5e-4) and shorter lifespan (rG = ‡0.21, SE = 0.09, P_FDR_ = 0.03). Finally, total psychopathy was associated with greater area-level socio-economic deprivation, as measured by the Townsend Index (rG = 0.54, SE = 0.14, P_FDR_ = 7.9e-4). Similar patterns of genetic correlations were observed for F1 and F2.

**Figure 4.**
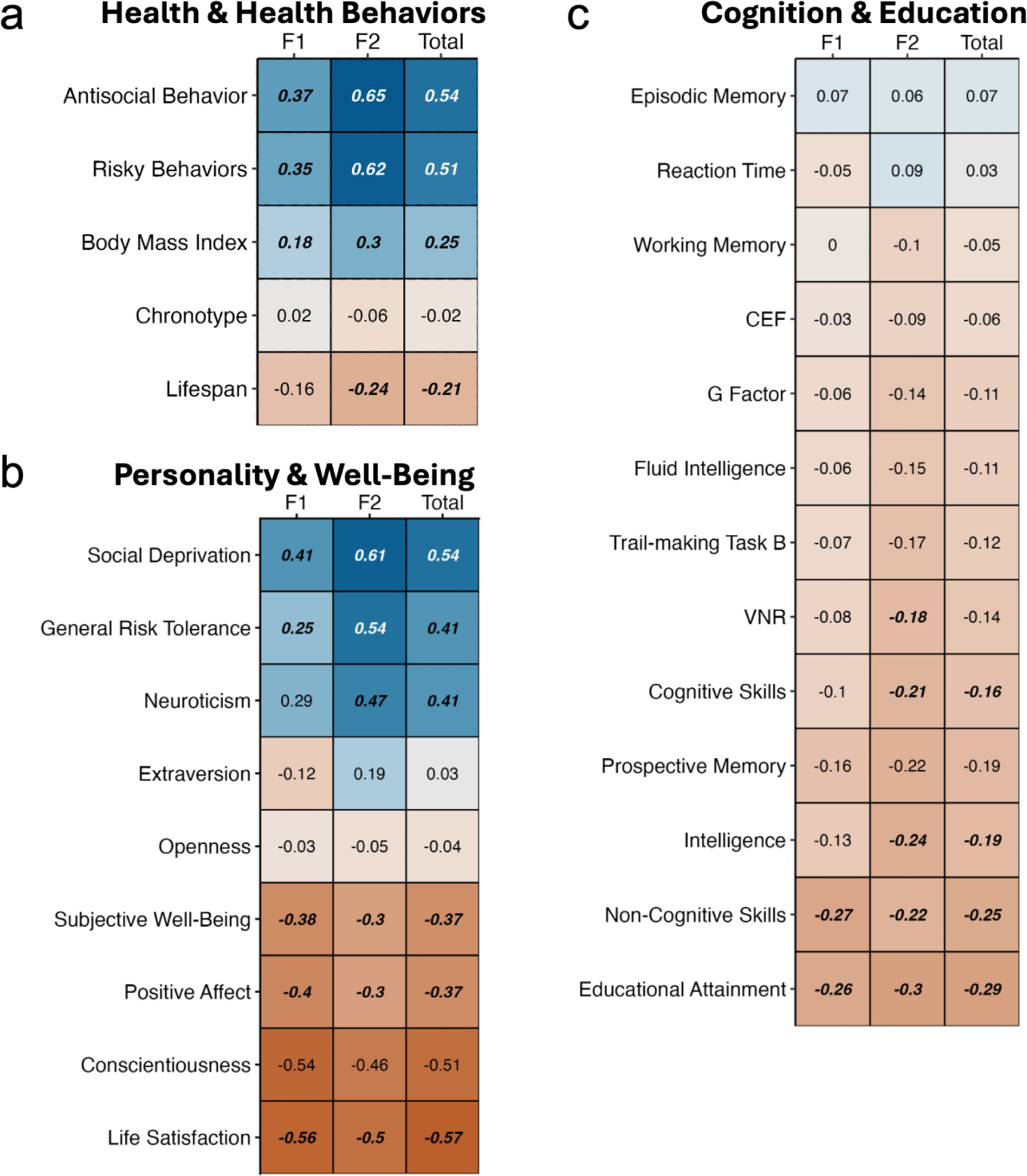
Psychopathy shows significant genetic overlap with health, personality, and cognition-related traits. Note. Heatmaps showing estimates of genetic correlation between self-reported psychopathy and external traits across three categories: Health Outcomes and Health-Related Behavior (panel a), Personality and Well-Being (panel b) and Cognition and Education (panel c). Estimates with false-discovery rate q-values less than 0.05 are indicated in bold and italics. CEF = Common Executive Function, VNR = Verbal-Numerical Reasoning.

#### Personality and Well-being

Personality and well-being traits also exhibited moderate genetic correlations with total psychopathy scores. Significant negative correlations were found with life satisfaction (rG = ‡0.57, SE = 0.19, P_FDR_ = 5.6e-3), positive affect (rG = ‡0.37, SE = 0.10, P_FDR_ = 7.9e-4), and subjective well-being (rG = ‡0.37, SE = 0.12, P_FDR_ = 5.3e-3). Total psychopathy scores showed significant positive genetic correlations with neuroticism (rG = 0.41, SE = 0.18, P_FDR_ = 0.03) and general risk tolerance (rG = 0.41, SE = 0.09, P_FDR_ = 2.3e-5). Genetic correlations with F1 and F2 were directionally consistent with those observed for the total score.

#### Cognition and Education

Finally, total psychopathy scores showed more modest genetic correlations with traits related to cognition and education. For most cognitive phenotypes, psychopathy scores exhibited non-significant genetic correlations, with nominal trends towards poorer cognitive performance. However, statistically significant genetic correlations were observed between total psychopathy scores and poorer cognitive (rG = ‡0.16, SE = 0.06, P_FDR_ = 0.02) and non-cognitive (rG = ‡0.25, SE = 0.07, P_FDR_ = 4.5e-3) skills, intelligence (rG = ‡0.19, SE = 0.06, P_FDR_ = 6.6e-3), as well as educational attainment (rG = ‡0.29, SE = 0.06, P_FDR_ = 8.0e-6). These significant associations were directionally consistent across F1 and F2. Only F2 showed a statistically significant genetic correlation with verbal-numerical reasoning (rG = ‡0.18, SE = 0.07, P_FDR_ = 1.3e-2), although F1 and total scores exhibited directionally consistent point estimates.

### Sensitivity Analysis

We performed a set of sensitivity analyses to compare the marginal and conditional genetic associations between SRP scores and external traits while controlling for case-control status. These suggested that genetic associations of SRP scores with external traits were largely independent of mood disorder case status (Supplementary Figure 2 and Supplementary Table 4). Given that these models are fully saturated (i.e. all degrees of freedom are used), it is difficult to formally test for a statistically significant difference between the marginal and conditional association estimates. However, marginal and conditional association estimates were highly correlated, with a Pearson correlation of 0.94 (95% CI: 0.91-0.96). Conditional estimates were attenuated on average compared to marginal associations, with the mean absolute difference between conditional and marginal estimates being 0.06 (95%CI: 0.05-0.08) for F1, 0.12 (95%CI: 0.09-0.15) for F2, and 0.09 (95%CI:0.07-0.11) for the total score.

## Discussion

In the largest genome-wide association study of psychopathy, we observed significant but modest common-variant heritability for total psychopathy scores and for the affective–interpersonal (F1) and antisocial–lifestyle (F2) subdimensions, with h^2^ estimates of ∼0.12–0.14. These values fall squarely within the range typically observed for psychiatric and behavioral phenotypes in large-scale GWAS(e.g. ^63–65)^ and are broadly consistent with twin and family studies indicating substantial genetic influence on psychopathic traits and callous–unemotional characteristics^1, 41,66,67^. Notably, the SNP-heritability for psychopathy in the present study appears substantially higher than that reported for broad antisocial behavior (∼0.03–0.08) in the largest meta-analyses of antisocial phenotypes^35,36^, and is comparable in magnitude to, or higher than, SNP-heritability estimates for several major psychiatric disorders such as major depression and ADHD^63,65^. Furthermore, the genetic correlation between psychopathy total scores and broad antisocial behavior was moderate (0.54) and significantly less than unity. This pattern supports the notion that psychopathy—operationalized as a dimensional trait—captures unique genetic variance over and above broadband antisocial or externalizing constructs.

Our finding that F1 and F2 show similar SNP-heritability and very high genetic correlations with each other and with the total psychopathy score is informative for ongoing debates about whether affective–interpersonal and antisocial–lifestyle features are etiologically distinct. Twin and family studies have generally reported moderate-to-high heritability for psychopathic traits in adults^40^ and callous-unemotional traits in children and adolescents^66,68^, with inconsistent evidence for strongly divergent genetic architectures across psychopathy trait factors^69,70^. The current data, with genetic correlations close to unity among F1, F2, and the total score (albeit with notably large standard errors), reinforce the view that these dimensions largely reflect a shared underlying polygenic liability rather than separate syndromes. At the same time, this does not preclude factor-specific neurocognitive mechanisms or contributions from rare variants^29^, but it suggests that such specificity is not sufficiently captured by common variants at presently available sample sizes. The absence of genome-wide significant loci, despite non-zero SNP-heritability, underscores the need for substantially larger samples in combination with multivariate approaches to achieve reliable locus discovery. Further, such approaches reduce the likelihood of Type-2 error by providing the power to detect real genetic associations when true rG is low, but non-zero.

At the cross-trait and transdiagnostic level, we found widespread positive genetic correlations between psychopathy scores and multiple psychiatric disorders, particularly borderline personality disorder, ADHD, major depressive disorder, post-traumatic stress disorder, autism spectrum disorder, schizophrenia, and substance use disorders. This pattern is broadly consistent with large cross-disorder analyses demonstrating pervasive sharing of common-variant risk across psychiatric conditions^64^, but extends this work by explicitly situating psychopathy within that network of genetic liabilities. The strong genetic overlap with borderline personality disorder, ADHD, and substance use disorders dovetails with externalizing-spectrum and HiTOP models that locate psychopathy—especially its antisocial–lifestyle component—within a broader disinhibitory/externalizing dimension^27,48,71^. Our observation that F2 appears to drive much of the genetic covariance with substance use disorders aligns with phenotypic data indicating that antisocial–lifestyle traits account for most of the overlap between psychopathy and externalizing outcomes^1,2^. Conversely, the absence of significant genetic correlation with compulsive disorders (obsessive–compulsive disorder, Tourette’s syndrome, anorexia nervosa) is consistent with evidence that “overcontrol” and “disinhibition” lie on partially dissociable higher-order dimensions^72–76^ and suggests that psychopathy’s genetic liabilities are more tightly coupled to disinhibition and negative affectivity than to overcontrolled or compulsive psychopathology. Importantly, we did not find evidence that F1 exerts a protective genetic effect on mood or anxiety disorders; instead, psychopathy scores as a whole showed positive genetic correlations with depression and PTSD, implying that any phenotypic resilience sometimes ascribed to “primary” psychopathy may not be reflected at the level of common-variant architecture in general adult samples. Future work using more extreme clinical phenotypes, and which characterize rare and structural variants are necessary to understand the degree to which these results are generalizable.

Beyond categorical psychiatric diagnoses, psychopathy exhibited substantial genetic covariance with health-related behaviors, somatic health, personality, well-being, and cognitive and educational outcomes. Positive genetic correlations with broad risky behavior and with a GWAS of broadband antisocial behavior indicate that our psychopathy phenotype indexes the same general liability toward rule-breaking, risk-taking, and externalizing behavior identified in large-scale genomic studies of externalizing and antisocial traits^35–37^. The positive genetic correlations with higher BMI and shorter lifespan are consistent with epidemiological findings linking psychopathic or antisocial traits to health-risk behaviors, injuries, chronic medical burden, and elevated all-cause and unnatural-cause mortality^1,17,18,77^. Likewise, the pattern of negative genetic correlations with life satisfaction, positive affect, and subjective well-being, and positive correlations with social deprivation, neuroticism and general risk tolerance, suggests that the common-variant architecture underlying psychopathy in this community sample is not affectively “neutral” or specifically characterized by low anxiety. Rather, in this community-based context it overlaps with genetic liabilities for emotional distress and reduced well-being, a profile more consistent with “secondary” or mixed psychopathy presentations than with a purely fearless, low-anxiety “primary” subtype^1,2,78^. Finally, modest but significant negative genetic correlations with cognitive and non-cognitive skills, intelligence, and educational attainment parallel the pattern observed for broad externalizing polygenic indices, which predict lower educational attainment, poorer cognitive performance, and extensive social and medical impairment^36,37,77^.

Taken together, these results reinforce several conceptual points. First, they are theoretically consistent with taxometric and longitudinal evidence that psychopathy is best conceptualized as a dimensional trait distributed across the population, rather than a discrete taxon. While we did not perform an empirical comparison of taxonomic versus dimensional liability models (e.g. GTACCC), the current results are compatible with a dimensional genetic architecture for psychopathy, characterized by modest, distributed polygenic signal and substantial—but incomplete—overlap with broader externalizing liability^1,2,4,21,79^. Second, they highlight that psychopathy—particularly when captured via dimensional scores in community-based samples—indexes not only risk for antisocial and substance use outcomes, but also a more global pattern of psychiatric, somatic, and functional burden, which aligns with recent public-health–oriented accounts^1,80,81^. Third, the somewhat higher SNP-heritability estimates for psychopathy relative to prior GWAS of broad antisocial behavior are consistent with the possibility that more narrowly defined, psychometrically coherent phenotypes may reduce heterogeneity and improve signal-to-noise for gene discovery. However, direct comparisons across studies must be interpreted cautiously given differences in phenotype definition, measurement, and ascertainment. Compelling inference requires an explicit test in future multi-phenotype and multivariate designs that harmonize measurement and analytic pipelines.^36,37^

Several limitations merit consideration. Despite robust SNP-heritability estimates and numerous significant genetic correlations, the sample remains underpowered for locus discovery in the context of a highly polygenic trait, as evidenced by the absence of genome-wide significant hits—an issue that has also constrained previous antisocial behavior and CU-trait GWAS^29,35,82^. As in most current GWAS, analyses were focused primarily on individuals of European ancestry, limiting generalizability and likely underestimating the contribution of ancestry-specific variants; larger multi-ancestry psychopathy GWAS will be crucial for improving generalizability and fine-mapping resolution^83^.

Further, we use variability in psychopathy trait scores in online survey respondents, rather than performing a clinical assessment of psychopathy in criminal offenders or forensic patients. Our psychopathy measure was designed as a self-report analog of Psychopathy Checklist - Revised (PCL-R) for measuring dimensional psychopathic traits in community samples^84,85^. However, as a self-report measure it may be influenced by recall or reporting bias and other related forms of measurement error. In addition, while the range of SRP-SF scores indicate that we sampled from each of the “low”, “average”, “elevated” and “extremely elevated” score bins (according to normed data from a community sample), mean scores for Total, F1 and F2 are all within the “average” score band^58^. Concern may thus be raised that we are not capturing the clinical/forensic construct of “psychopathy,” which is often operationalized as a score of 30 or above on the PCL-R. However, the use of a dimensional measure of psychopathy outside of a correctional or forensic setting is defensible given strong evidence above in favor of a dimensional structure of psychopathy in community samples generally^86^ and those measured by the SRP-SF in particular^84^. Moreover, while mean SRP-SF values are higher in criminal offender vs. community populations, the distribution of psychopathy traits in criminal offenders overlaps community-level personality variation ^58,84^.

However, we note that the current study does not include individuals with extreme SRP scores, likely due to our method of recruitment and phenotype ascertainment for the AFFECT study. This restriction in range constitutes a limitation that may have artifactually reduced our heritability estimates and limited our ability to detect genetic correlations. Likewise, this measure was given at the last study timepoint, with a response rate of 28% (comparable to response rates of other online mental health assessments, such as those administered in the UK Biobank)^87^. This may have oversampled more conscientious individuals^87^, who would be more likely to complete the study, potentially further reducing the magnitude of our genetic signal for psychopathy.

Within these constraints, the present study indicates that psychopathy is a moderately heritable, highly polygenic trait with pleiotropic connections to externalizing, internalizing, health-risk behaviors, and adverse health and functional outcomes. Rather than representing a genetically isolated “forensic” construct, psychopathy appears to sit at the intersection of broad transdiagnostic dimensions of psychopathology and health, with implications for its nosological status, for prevention, and for intervention strategies.

## Data Availability

Data produced in this study are available on request to the 23andMe Research Institute

## Acknowledgements

J.D.T. is supported by NIH grant K01MH141330. T.T.M. is supported by NIH grant K08MH135343. The AFFECT study was funded by H. Lundbeck A/S and the Milken Institute.

We would like to thank the research participants and employees of 23andMe Research Institute for making this work possible.

23andMe Research Team: Stella Aslibekyan, Adam Auton, Robert K. Bell, Katelyn Kukar Bond, Zayn Cochinwala, Sayantan Das, Kahsaia de Brito, Emily DelloRusso, Chris Eijsbouts, Sarah L. Elson, Chris German, Julie M. Granka, Barry Hicks, David A. Hinds, Reza Jabal, Aly Khan, Matthew J. Kmiecik, Alan Kwong, Yanyu Liang, Keng-Han Lin, Matthew H. McIntyre, Shubham Saini, Anjali J. Shastri, Jingchunzi Shi, Suyash Shringarpure, Qiaojuan Jane Su, Vinh Tran, Joyce Y. Tung, Catherine H. Weldon, Wanwan Xu

## Supplementary Information

**Supplementary Figure 1.**
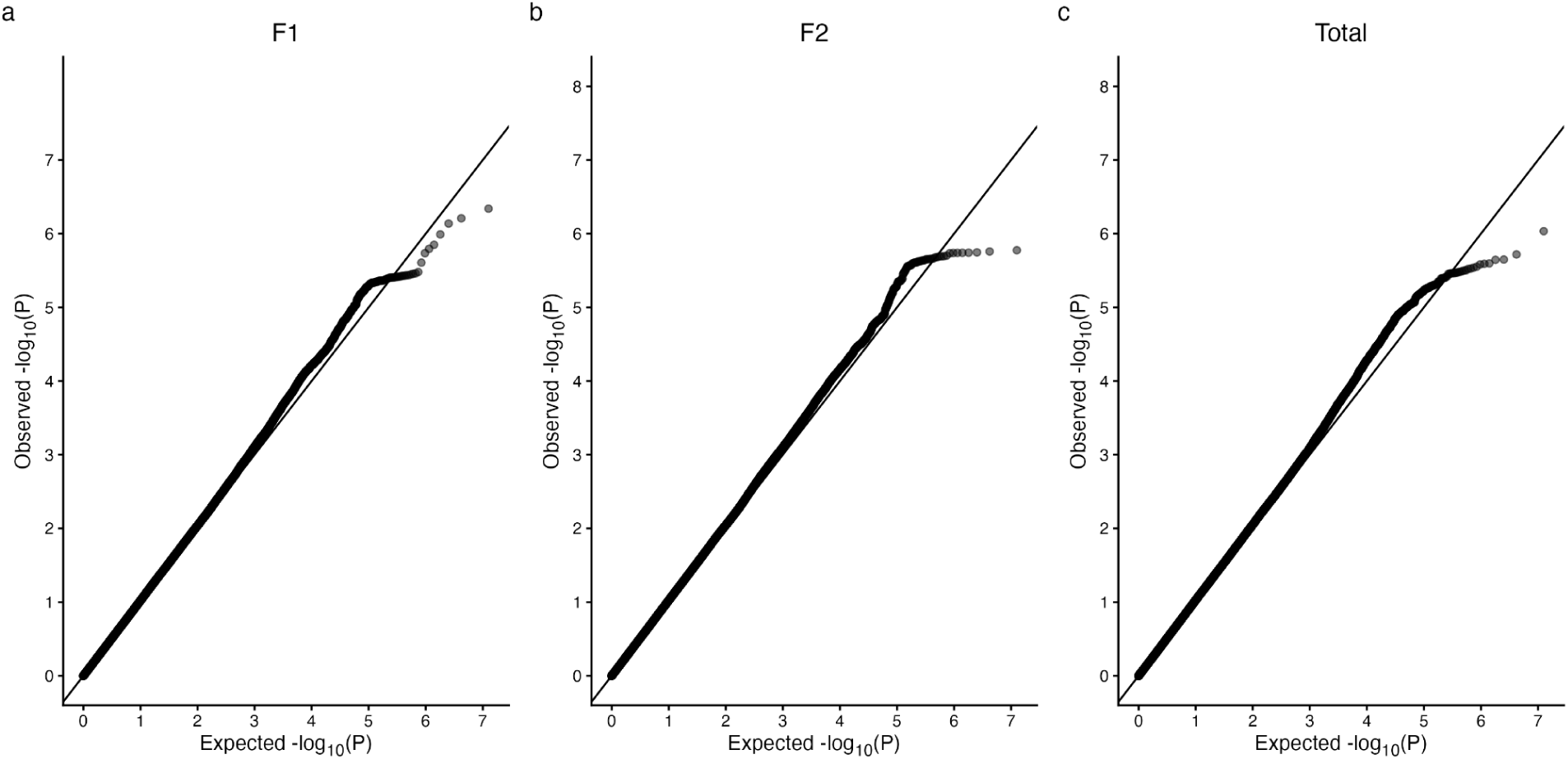
GWAS Quantile Quantile Plots. Note. Quantile-quantile plots show the observed (y axis) versus expected (x axis) -log_10_p-values from genome-wide association studies of self-reported psychopathy factor F1 (panel a), F2 (panel b), and total (panel c) scores.

**Supplementary Figure 2.**
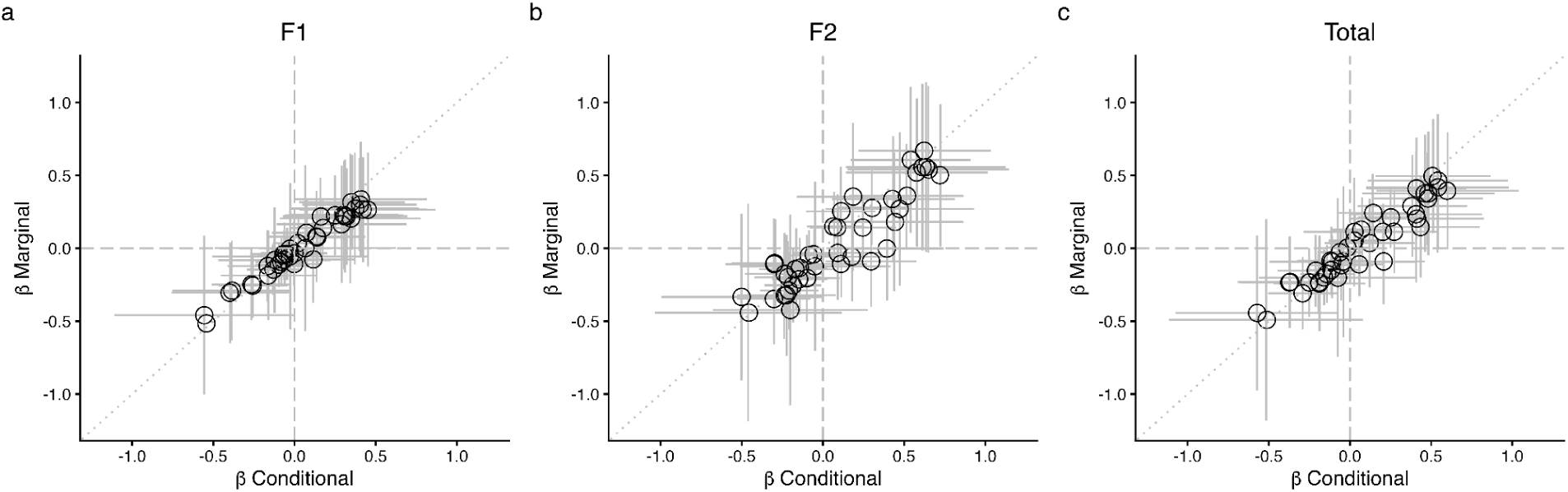
Marginal vs Conditional Genetic Regression Coefficient Estimates. Note. As a sensitivity analysis, GenomicSEM was used to estimate regression coefficients of each external trait on each self-reported psychopathy trait. Marginal estimates are shown on the y axis, and conditional coefficient estimates (controlling for a within-sample GWAS of mood disorder case-control status) are shown on the x axis. The dotted line depicts a null expectation of intercept 0,0 and slope 1.

## References

1. De Brito, S. A. D. et al. Psychopathy. Nat. Rev. Dis. Primers 7, 49 (2021).

2. Patrick, C. J. Psychopathy: Current Knowledge and Future Directions. Annu. Rev. Clin. Psychol. 18, 387–415 (2022).

3. Hare, R. D. & Neumann, C. S. Psychopathy as a Clinical and Empirical Construct. Annu. Rev. Clin. Psychol. 4, 217–246 (2008).

4. Skeem, J. L., Polaschek, D. L. L., Patrick, C. J. & Lilienfeld, S. O. Psychopathic Personality. Psychol. Sci. Public Interest 12, 95–162 (2011).

5. Weaver, S. S., Dargis, M., Kiehl, K. A. & Koenigs, M. Criminal histories and rates of recidivism among two subtypes of psychopathic individuals. Crim. Justice Behav. 49, 471–491 (2022).

6. Hemphill, J. F., Hare, R. D. & Wong, S. Psychopathy and recidivism: A review. Legal Criminol. Psychol. 3, 139–170 (1998).

7. Braga, T., de Castro Rodrigues, A., Cruz, A. R., Pechorro, P. & Cunha, O. The four facets of the Psychopathy Checklist, Youth Version and recidivism: A meta-analysis. Aggress. Violent Behav. 70, 101824 (2023).

8. Allen, C. H. et al. Psychopathy scores predict recidivism in high-risk youth: A five-year follow-up study. Res. Child Adolesc. Psychopathol. 52, 1089–1103 (2024).

9. Edens, J. F., Cox, J., Smith, S. T., DeMatteo, D. & Sörman, K. How reliable are Psychopathy Checklist-Revised scores in Canadian criminal trials? A case law review. Psychol. Assess. 27, 447–456 (2015).

10. DeMatteo, D. et al. Statement of concerned experts on the use of the Hare Psychopathy Checklist—Revised in capital sentencing to assess risk for institutional violence. Psychol. Public Policy Law 26, 133–144 (2020).

11. Morse, S. J. Preventive detention of psychopaths and dangerous offenders. Handbook on psychopathy and law (2013).

12. Schopp, R. F. & Slain, A. J. Psychopathy, criminal responsibility, and civil commitment as a sexual predator. Behav. Sci. Law 18, 247–274 (2000).

13. Holper, L. et al. Criterion validity of the Psychopathy Checklist in legal contexts: An updated meta-analysis. J. Pers. Assess. 107, 547–562 (2025).

14. Anderson, D. A. The Aggregate Cost of Crime in the United States. J. Law Econ. 64, 857–885 (2021).

15. Gatner, D. T., Douglas, K. S., Almond, M. F. E., Hart, S. D. & Kropp, P. R. How much does that cost? Examining the economic costs of crime in North America attributable to people with psychopathic personality disorder. Personal. Disord. 14, 391–400 (2023).

16. Shadid, J. et al. The global epidemiology of personality disorder: a systematic review and meta-regression. Lancet Psychiatry 12, 932–946 (2025).

17. Maurer, J. M. et al. Adolescents with elevated psychopathic traits are associated with an increased risk for premature mortality. Res. Child Adolesc. Psychopathol. 53, 17–28 (2025).

18. Vaurio, O., Repo-Tiihonen, E., Kautiainen, H. & Tiihonen, J. Psychopathy and mortality. J. Forensic Sci. 63, 474–477 (2018).

19. Seara-Cardoso, A. & Viding, E. Functional Neuroscience of Psychopathic Personality in Adults. J. Pers. 83, 723–737 (2015).

20. Coid, J. & Yang, M. The distribution of psychopathy among a household population: categorical or dimensional? Soc. Psychiatry Psychiatr. Epidemiol. 43, 773–781 (2008).

21. Edens, J. F., Marcus, D. K., Lilienfeld, S. O. & Poythress, N. G., Jr. Psychopathic, not psychopath: taxometric evidence for the dimensional structure of psychopathy. J. Abnorm. Psychol. 115, 131–144 (2006).

22. Guay, J.-P., Ruscio, J., Knight, R. A. & Hare, R. D. A taxometric analysis of the latent structure of psychopathy: evidence for dimensionality. J. Abnorm. Psychol. 116, 701–716 (2007).

23. Gaule, A. et al. Reduced prosocial motivation and effort in adolescents with conduct problems and callous-unemotional traits. J. Child Psychol. Psychiatry 65, 1061–1071 (2024).

24. Carlisi, C. O. et al. Differential mapping of psychopathic traits and general psychopathology in a large young adult sample. J. Pers. Disord. 38, 535–558 (2024).

25. Phillips, N. L., Rose, L., Lynam, D. R. & Miller, J. D. Elemental psychopathy assessment’s nomological net: A meta-analytic review. Personal. Disord. 16, 377–393 (2025).

26. Hildebrand, M. & de Ruiter, C. PCL-R psychopathy and its relation to DSM-IV Axis I and II disorders in a sample of male forensic psychiatric patients in The Netherlands. Int. J. Law Psychiatry 27, 233–248 (2004).

27. Krueger, R. F. et al. Validity and utility of Hierarchical Taxonomy of Psychopathology (HiTOP): II. Externalizing superspectrum. World Psychiatry 20, 171–193 (2021).

28. Patrick, C. J., Hicks, B. M., Krueger, R. F. & Lang, A. R. Relations between psychopathy facets and externalizing in a criminal offender sample. J. Pers. Disord. 19, 339–356 (2005).

29. Pezzoli, P., McCrory, E. J. & Viding, E. Shedding light on antisocial behavior through genetically informed research. Annu. Rev. Psychol. 76, 797–819 (2025).

30. Jones, A. P., Laurens, K. R., Herba, C. M., Barker, G. J. & Viding, E. Amygdala hypoactivity to fearful faces in boys with conduct problems and callous-unemotional traits. Am. J. Psychiatry 166, 95–102 (2009).

31. Hicks, B. M., Krueger, R. F., Iacono, W. G., McGue, M. & Patrick, C. J. Family transmission and heritability of externalizing disorders: a twin-family study: A twin-family study. Arch. Gen. Psychiatry 61, 922–928 (2004).

32. Hicks, B. M., Foster, K. T., Iacono, W. G. & McGue, M. Genetic and environmental influences on the familial transmission of externalizing disorders in adoptive and twin offspring. JAMA Psychiatry 70, 1076–1083 (2013).

33. Gustavson, D. E., Franz, C. E., Panizzon, M. S., Lyons, M. J. & Kremen, W. S. Internalizing and externalizing psychopathology in middle age: genetic and environmental architecture and stability of symptoms over 15 to 20 years. Psychol. Med. 50, 1530–1538 (2020).

34. Kendler, K. S. et al. A Swedish population-based multivariate twin study of externalizing disorders. Behav. Genet. 46, 183–192 (2016).

35. Tielbeek, J. J. et al. Genome-Wide Association Studies of a Broad Spectrum of Antisocial Behavior. JAMA Psychiatry 74, 1242 (2017).

36. Tielbeek, J. J. et al. Uncovering the genetic architecture of broad antisocial behavior through a genome-wide association study meta-analysis. Mol. Psychiatry 27, 4453–4463 (2022).

37. Linnér, R. K. et al. Multivariate analysis of 1.5 million people identifies genetic associations with traits related to self-regulation and addiction. Nat. Neurosci. 24, 1367–1376 (2021).

38. Barr, P. B. et al. Parsing genetically influenced risk pathways: genetic loci impact problematic alcohol use via externalizing and specific risk. Transl. Psychiatry 12, 420 (2022).

39. Viding, E., Blair, R. J. R., Moffitt, T. E. & Plomin, R. Evidence for substantial genetic risk for psychopathy in 7-year-olds. J. Child Psychol. Psychiatry 46, 592–597 (2005).

40. Dotterer, H. L. et al. Elucidating the role of negative parenting in the genetic v. environmental influences on adult psychopathic traits. Psychol. Med. 53, 897–907 (2023).

41. Moore, A. A., Blair, R. J., Hettema, J. M. & Roberson-Nay, R. The genetic underpinnings of callous-unemotional traits: A systematic research review. Neurosci. Biobehav. Rev. 100, 85–97 (2019).

42. Dhanani, S. et al. A systematic review of the heritability of specific psychopathic traits using Hare’s two-factor model of psychopathy. CNS Spectr. 23, 29–38 (2018).

43. Buckholtz, J. W., Karmarkar, U., Ye, S., Brennan, G. M. & Baskin-Sommers, A. Blunted ambiguity aversion during cost-benefit decisions in antisocial individuals. Sci. Rep. 7, 2030 (2017).

44. Baskin-Sommers, A., Stuppy-Sullivan, A. M. & Buckholtz, J. W. Psychopathic individuals exhibit but do not avoid regret during counterfactual decision making. Proc. Natl. Acad. Sci. U. S. A. 113, 14438–14443 (2016).

45. Hosking, J. G. et al. Disrupted Prefrontal Regulation of Striatal Subjective Value Signals in Psychopathy. Neuron 95, 221–231.e4 (2017).

46. Rodman, A. M. et al. Selective Mapping of Psychopathy and Externalizing to Dissociable Circuits for Inhibitory Self-Control. Clin. Psychol. Sci. 4, 559–571 (2015).

47. Krueger, R. F., Markon, K. E., Patrick, C. J., Benning, S. D. & Kramer, M. D. Linking antisocial behavior, substance use, and personality: an integrative quantitative model of the adult externalizing spectrum. J. Abnorm. Psychol. 116, 645–666 (2007).

48. Rodrik, O., Weaver, S. S., Kiehl, K. A. & Koenigs, M. Correlates of Externalizing Psychopathology in Incarcerated Men. Psychol. Assess. 34, 912–922 (2022).

49. Sellbom, M. Elucidating the validity of the externalizing spectrum of psychopathology in correctional, forensic, and community samples. J. Abnorm. Psychol. 125, 1027–1038 (2016).

50. Coid, J. et al. Psychopathy among prisoners in England and Wales. Int. J. Law Psychiatry 32, 134–141 (2009).

51. Widiger, T. & Crego, C. Handbook Psychopathy 2nd Edn. 281–298 (Guilford Press, Patrick, C. J., 2018).

52. Hicks, B. M. & Drislane, L. E. Variants (‘subtypes’) of psychopathy. Handbook of psychopathy 2, 297–333 (2018).

53. Blonigen, D. M., Hicks, B. M., Krueger, R. F., Patrick, C. J. & Iacono, W. G. Psychopathic personality traits: heritability and genetic overlap with internalizing and externalizing psychopathology. Psychol. Med. 35, 637–648 (2005).

54. Buckholtz, J. W. Social norms, self-control, and the value of antisocial behavior. Curr. Opin. Behav. Sci. 3, 122–129 (2015).

55. Karlsson Linnér, R., et al. Genome-wide association analyses of risk tolerance and risky behaviors in over 1 million individuals identify hundreds of loci and shared genetic influences. Nat. Genet. 51, 245–257 (2019).

56. Dalby, M. et al. Characterizing mood disorders in the AFFECT study: a large, longitudinal, and phenotypically rich genetic cohort in the US. Transl. Psychiatry 12, 121 (2022).

57. Neumann, C. S. & Pardini, D. Factor structure and construct validity of the Self-Report Psychopathy (SRP) scale and the Youth Psychopathic Traits Inventory (YPI) in young men. J. Pers. Disord. 28, 419–433 (2014).

58. Delroy L. Paulhus, Craig S. Neumann, Robert D. Hare. Self-Report Psychopathy Scale-Fourth Edition. (2016).

59. Henn, B. M. et al. Cryptic distant relatives are common in both isolated and cosmopolitan genetic samples. PLoS One 7, e34267 (2012).

60. Bulik-Sullivan, B. et al. LD Score regression distinguishes confounding from polygenicity in genome-wide association studies. Nat. Genet. 47, 291–295 (2015).

61. Bulik-Sullivan, B. et al. An Atlas of Genetic Correlations across Human Diseases and Traits. Nat. Genet. 47, 1236–1241 (2015).

62. Grotzinger, A. D., et al. GenomicSEM: Structural Equation Modeling Based on GWAS Summary Statistics.

63. Major Depressive Disorder Working Group of the Psychiatric Genomics Consortium. Electronic address: & Major Depressive Disorder Working Group of the Psychiatric Genomics Consortium. Trans-ancestry genome-wide study of depression identifies 697 associations implicating cell types and pharmacotherapies. Cell 188, 640–652.e9 (2025).

64. Grotzinger, A. D. et al. Mapping the genetic landscape across 14 psychiatric disorders. Nature (2025) doi:10.1038/s41586-025-09820-3.

65. van der Laan, C. M. et al. Genome-wide association meta-analysis of childhood ADHD symptoms and diagnosis identifies new loci and potential effector genes. Nat. Genet. 57, 2427–2435 (2025).

66. Takahashi, Y., Pease, C. R., Pingault, J.-B. & Viding, E. Genetic and environmental influences on the developmental trajectory of callous-unemotional traits from childhood to adolescence. J. Child Psychol. Psychiatry 62, 414–423 (2021).

67. Hung, I.-T., Viding, E., Stringaris, A., Ganiban, J. M. & Saudino, K. J. Understanding the etiology of Externalizing Problems in Young Children: The Roles of callous-Unemotional Traits and irritability. J. Am. Acad. Child Adolesc. Psychiatry 64, 1420–1433 (2025).

68. Tomlinson, R. C., Hyde, L. W., Dotterer, H. L., Klump, K. L. & Burt, S. A. Parenting moderates the etiology of callous-unemotional traits in middle childhood. J. Child Psychol. Psychiatry 63, 912–920 (2022).

69. Larsson, H., Andershed, H. & Lichtenstein, P. A genetic factor explains most of the variation in the psychopathic personality. J. Abnorm. Psychol. 115, 221–230 (2006).

70. Taylor, J., Loney, B. R., Bobadilla, L., Iacono, W. G. & McGue, M. Genetic and environmental influences on psychopathy trait dimensions in a community sample of male twins. J. Abnorm. Child Psychol. 31, 633–645 (2003).

71. Davis, C. N. et al. Integrating HiTOP and RDoC frameworks Part I: Genetic architecture of externalizing and internalizing psychopathology. Psychol. Med. 55, e138 (2025).

72. Christian, C. et al. Latent profile analysis of impulsivity and perfectionism dimensions and associations with psychiatric symptoms. J. Affect. Disord. 283, 293–301 (2021).

73. Gilbert, K. et al. Thin slice derived personality types predict longitudinal symptom trajectories. Personal. Disord. 12, 275–285 (2021).

74. Gilmartin, T., Dipnall, J. F., Gurvich, C. & Sharp, G. Identifying overcontrol and undercontrol personality types among young people using the five factor model, and the relationship with disordered eating behaviour, anxiety and depression. J. Eat. Disord. 12, 16 (2024).

75. Gilmartin, T., Gurvich, C., Dipnall, J. F. & Sharp, G. Using the alternative model of personality disorders for DSM-5 traits to identify personality types, and the relationship with disordered eating, depression, anxiety and stress. J. Eat. Disord. 13, 19 (2025).

76. Steinhoff, M. F. et al. Childhood behavioral inhibition and overcontrol: Risk for psychiatric and peer outcomes. Dev. Psychol. (2025) doi:10.1037/dev0002042.

77. Tielbeek, J. J. & Boutwell, B. B. Exploring the genomic architectures of health, physical traits and antisocial behavioral outcomes: A brief report. Front. Psychiatry 11, 539 (2020).

78. Sethi, A. et al. Primary and Secondary Variants of Psychopathy in a Volunteer Sample Are Associated With Different Neurocognitive Mechanisms. Biol. Psychiatry Cogn. Neurosci. Neuroimaging 3, 1013–1021 (2018).

79. Sica, C. et al. Comparing the DSM-5 dimensional trait and triarchic model conceptions of psychopathy: An external validity analysis. J. Pers. Disord. 38, 368–400 (2024).

80. Reidy, D. E. & Bogen, K. W. Public health considerations in psychopathy. in Dangerous Behavior in Clinical and Forensic Psychology 611–635 (Springer International Publishing, Cham, 2022).

81. Viding, E., McCrory, E., Baskin-Sommers, A., De Brito, S. & Frick, P. An ‘embedded brain’ approach to understanding antisocial behaviour. Trends Cogn. Sci. 28, 159–171 (2024).

82. Viding, E. et al. In search of genes associated with risk for psychopathic tendencies in children: a two-stage genome-wide association study of pooled DNA: Genome-wide association study of psychopathic tendencies. J. Child Psychol. Psychiatry 51, 780–788 (2010).

83. Martin, A. R. et al. Clinical use of current polygenic risk scores may exacerbate health disparities. Nature Genetics 2019 51:4 51, 584–591 (2019).

84. Dotterer, H. L. et al. Examining the factor structure of the self-Report of psychopathy Short-Form across four young adult samples. Assessment 24, 1062–1079 (2017).

85. Thomson, N. D., Kjaervik, S. L., Neumann, C. S. & Hare, R. D. The role of psychopathy in subtypes of aggression and gun violence. J. Psychopathol. Behav. Assess. 47, 69 (2025).

86. Neumann, C. S. & Hare, R. D. Psychopathic traits in a large community sample: links to violence, alcohol use, and intelligence. J. Consult. Clin. Psychol. 76, 893–899 (2008).

87. Davis, K. A. S. et al. Mental health in UK Biobank - development, implementation and results from an online questionnaire completed by 157 366 participants: a reanalysis. BJPsych Open 6, e18 (2020).

